# Exploring diseases/traits and blood proteins causally related to expression of ACE2, the putative receptor of SARS-CoV-2: A Mendelian Randomization analysis highlights tentative relevance of diabetes-related traits

**DOI:** 10.1101/2020.03.04.20031237

**Authors:** Shitao Rao, Alexandria Lau, Hon-Cheong So

## Abstract

**Objectives:** COVID-19 has become a major public health problem. There is good evidence that ACE2 is a receptor for SARS-CoV-2, and high expression of *ACE2* may increase susceptibility to infection. We aimed to explore risk factors affecting susceptibility to infection and prioritize drug repositioning candidates, based on Mendelian randomization (MR) studies on *ACE2* lung expression.

**Methods:** We conducted a phenome-wide MR study to prioritize diseases/traits and blood proteins causally linked to *ACE2* lung expression in GTEx. We also explored drug candidates whose targets overlapped with the top-ranked proteins in MR, as these drugs may alter *ACE2* expression and may be clinically relevant.

**Results:** The most consistent finding was tentative evidence of an association between diabetes-related traits and increased *ACE2* expression. Based on one of the largest GWAS on type 2 diabetes (T2DM) to date (*N*=898,130), T2DM was causally linked to raised *ACE2* expression(p=2.91E-03;MR-IVW). Significant associations(at nominal level; *p*<0.05) with *ACE2* expression was observed across multiple DM datasets and analytic methods, for type 1 and 2 diabetes and related traits including early start of insulin. Other diseases/traits having nominal significant associations with increased expression included inflammatory bowel disease, (ER+)breast and lung cancers, asthma, smoking and elevated ALT. We also identified drugs that may target the top-ranked proteins in MR, such as fostamatinib and zinc.

**Conclusions:** Our analysis suggested that diabetes and related traits may increase *ACE2* expression, which may influence susceptibility to infection (or more severe infection). However, none of these findings withstood rigorous multiple testing corrections (at FDR<0.05). Proteome-wide MR analyses might help uncover mechanisms underlying *ACE2* expression and guide drug repositioning. Further studies are required to verify our findings.

## Introduction

Coronavirus Disease 2019 (COVID-19), caused by SARS-CoV-2, has resulted a pandemic affecting more than a hundred countries worldwide (1-3). More than 2 million confirmed cases have been reported worldwide as at 22 Apr 2020 (https://www.who.int/emergencies/diseases/novel-coronavirus-2019/situation-reports), while many mild or asymptomatic cases may remain undetected. Considering the severity of the outbreak, it is urgent to seek solutions to control the spread of the disease to susceptible groups, and to identify effective treatments. A better understanding of its pathophysiology is also urgently needed.

Notably, recent studies showed that over one quarter of confirmed cases had a history of comorbid conditions, such as hypertension, diabetes, cardiovascular disease and respiratory diseases (Table S1) (2-4). In addition, the severity of disease is likely be higher in patients with chronic conditions(2). However, it is unclear whether such comorbidities are *causally* related to increased susceptibility, and if so, what the underlying mechanisms may be. Confounding bias (e.g. by age, sex, comorbidities, medications received, smoking/drinking history etc.) may lead to spurious associations that preclude conclusions about causality. Establishing causality is important as this is closely related to the effectiveness of interventions. If a risk factor is *causally* related to an outcome, then interventions on the risk factor will lead to reduced risks of the outcome, which may not be true for associations per se.

Based on analysis of potential receptor usage and the released sequences of SARS-CoV-2, Wan *et al*. proposed that the host receptor of SARS-CoV-2 is angiotensin-converting enzyme 2 (ACE2). Virus infectivity studies on HELA cell lines further confirmed that ACE2 is a cellular entry receptor for SARS-CoV-2 (8). Another line of evidence came from structural study of SARS-CoV-2. Wrapp *et al*. observed that the ACE2 protein could bind to the SARS-CoV-2 spike ecto-domain with high affinity (9). Importantly, ACE2 has previously been established a receptor for SARS-CoV (6; 7). Taken together, the above provide strong evidence that ACE2 is a key receptor of the novel coronavirus.

A number of studies have looked into the relationship between ACE2 expression level and coronavirus infection. For example, it was found that overexpression of ACE2 protein lead to more efficient SARS-CoV replication, which was blocked by anti-ACE2 antibodies in a dose-dependent manner (6). Two further studies also showed that susceptibility to SARS-CoV infection was correlated with ACE2 expression in cell lines (10) (11). It is therefore reasonable to hypothesize that ACE expression also affects susceptibility to SARS-CoV-2 infection. Revealing diseases/traits causally associated with altered ACE2 expression may shed light on why certain individuals are more susceptible to SARS-CoV-2 infection (or more severe infections) and the underlying mechanisms (whether the increased susceptibility is mediated via ACE2).

In this study, we wish to answer the following question: what conditions or traits may lead to increased *ACE2* expression, which may in turn result in higher susceptibility to SARS-CoV-2 infection? Here we conducted a phenome-wide Mendelian randomization (MR) study to explore diseases/traits that may be *causally* linked to increased *ACE2* lung expression. Our study is *different* from most existing MR studies: instead of considering a disease as outcome, the outcome measure is *ACE2* expression, interpreted as a surrogate for susceptibility to infection, and the exposures tested are diseases/traits. While a number of tissues may also be affected by SARS-CoV-2(12), pneumonia is a common and major complication of the disease (3), hence we focused on lung expression in this study. Regarding our study approach, phenome-wide MR is a data-driven approach which has been performed in other contexts as a powerful way to uncover unknown causal risk factors for diseases (13-15). This approach allows multiple risk factors or outcomes to be studied simultaneously. MR makes use of genetic variants as “instruments” to represent the exposure of interest, and infers causal relationship between the exposure and outcome (16). In general, MR is not affected by reverse causality (17), as genetic variants are fixed at conception (which precedes the outcome). MR is also less susceptible to confounding bias compared to conventional case-control/cohort studies, as genetic instruments are usually less strongly associated with environmental exposures than ordinary risk factors(18) (please also refer to Supplementary Text for more detailed descriptions).

In addition to diseases, as a secondary analysis, we also studied serum/plasma proteins as exposure, as they may point to potential molecular mechanisms underlying *ACE2* expression and may serve as potential predictive or prognostic biomarkers. Such proteome-wide studies may help to reveal drug repositioning candidates (19), through the search for drugs that target the top-ranked proteins. For example, if a protein causally increases the risk of a disease, then by the definition of causality, blocking the protein will lead to reduced disease risks. By finding plasma/serum proteins *causally* linked to *ACE2* expression, one may find drugs altering *ACE2* expression, which in turn may be useful for treatment.

## Methods

### Genome-wide association study (GWAS) data

All GWAS data are extracted from publicly available databases, which are detailed below.

#### Exposure data

Most GWAS data employed were based on predominantly European samples, and proper correction for population stratification has been performed. Please also refer to Table S2a/2b for details on the ethnic composition and methods to account for population stratification for GWAS included in this work.

To perform the phenome-wide study, here we made use of the latest IEU (<u>I</u>ntegrative <u>E</u>pidemiology <u>U</u>nit of University of Bristol) GWAS database (https://gwas.mrcieu.ac.uk/), which contains up to 111,908,636,549 genetic associations from 31,773 GWAS summary datasets (as at 26^th^ Feb 2020). Details of each GWAS study may be retrieved from https://gwas.mrcieu.ac.uk/datasets/. The database was retrieved *via* the R package “TwoSampleMR” (ver 0.5.1). MR analysis was conducted with the same package. Due to the extremely huge number of traits in the database, we performed some pre-selection to the list of traits/diseases before full analysis.

Briefly, we selected the following categories of traits: **(1)** Traits listed as priority 1 (high priority) and labelled as “Disease” or “Risk factor” (81 and 71 items respectively); **(2)** traits labelled as “protein” [3371 items originally studied in ref(20; 21)]; **(3)** (selected) traits from the UK Biobank (UKBB), as it is one of the largest source of GWAS data worldwide (*N*~500,000). We consider that a proportion of traits have presumably low prior probability of association with respiratory infections, and others are less directly clinically relevant. To reduce computational burden and for ease of interpretation, a proportion of UKBB traits were filtered. More specifically, we excluded GWAS data of diseases or traits related to the following: eye or hearing problems, orthopedic and trauma-related conditions (except autoimmune diseases), skin problems (except systemic or autoimmune diseases), perinatal and obstetric problems, operation history, medication history (as confounding by indication is common and may affect the validity of results (22)), diet/exercise habit (as accuracy of information cannot be fully guaranteed and recall bias may be present), other socioeconomic features (such as type of jobs). A total of 425 UKBB traits were retained for final analysis under the third category. GWAS of blood proteins and UKBB traits were restricted to European samples.

GWAS of UKBB were based on analysis results from the Neale Lab (https://sites.google.com/broadinstitute.org/ukbbgwasresults/) and from MRC-IEU. GWAS analysis was performed using linear models with adjustment for population stratification; details of the analytic approach are given in: https://github.com/Nealelab/UK_Biobank_GWAS/tree/master/imputed-v2-gwas, http://www.nealelab.is/blog/2017/9/11/details-and-considerations-of-the-uk-biobank-gwas and https://doi.org/10.5523/bris.pnoat8cxo0u52p6ynfaekeigi. For binary outcomes, we converted the regression coefficients obtained from the linear model to those under a logistic model, based on methodology presented in (23). The SE under a logistic model was derived by the delta method (equation 37 in (23)).

#### Outcome data

The outcome was pulmonary expression of *ACE2*. While ideally one should study the protein expression in the lung, such data is scarce and corresponding genotype data (required for MR) is not available. Here we focus on the gene expression of *ACE2* in the lung (*N*=515). We retrieved GWAS summary data from the Genotype-Tissue Expression (GTEx) database (with API); it is one of the largest databases to date with both genotype and expression data for a large variety of tissues. The majority of the GTEx samples are European in ancestry (~85%); other ancestries included African Americans, Asians and American Indians(Table S2a). Population stratification was controlled by inclusion of principal components in genetic association analysis. For further details of GTEx please refer to (24); the eQTL analysis procedure is described in https://gtexportal.org/home/documentationPage#staticTextAnalysisMethods.

### Mendelian randomization (MR) analysis

Here we performed two-sample MR, in which the instrument-exposure and instrument-outcome associations were estimated in different samples.

#### Instrument SNP selection

MR was performed on (approximately) independent SNPs with r^2^ threshold of 0.001, following default settings in the R package TwoSampleMR. SNPs passing genome-wide significance (*p*<5e-8) were included as instruments. Clinical traits or blood proteins were treated as exposures, and we used the ‘extract_instruments’ function in TwoSampleMR to retrieve SNPs for each trait from corresponding GWAS. The source GWAS for each exposure are listed in Table S2. Only SNPs with available SNP-exposure and SNP-outcome association data were retained.

#### MR methods

We conducted MR primarily with the ‘inverse-variance weighted’ (MR-IVW) (25) and Egger regression (MR-Egger) (26) approaches, which are among the most widely used MR methods. For exposure with only one instrument, the Wald ratio method was used. For analysis with <3 genetic instruments, we employed MR-IVW only since MR-Egger cannot be reliably performed. The intercept from MR-Egger was used to evaluate presence of significant directional (imbalanced) horizontal pleiotropy.

For selected traits with at least nominally significant associations by MR-IVW or MR-Egger (p<0.05), we also performed further analysis by GSMR (Generalised Summary-data-based Mendelian Randomisation), weighted median (an ‘implicit’ outlier-removal method (30)) and MR-RAPS. GSMR also accounts for correlated SNPs and removes likely pleiotropic outliers (27).

We tried several r^2^ thresholds (0.001, 0.05, 0.1, 0.15, 0.2) for GSMR analysis on diabetes based on Mahajan et al.(28) (see Results/Table 2). SNP correlations were derived from 1000-Genomes European samples. MR-RAPS(29) is another methodology which takes into account multiple weak instruments by a robust procedure; we employed a more relaxed p-value threshold for SNP selection (0.01) for this method. One of the major concerns of MR is horizontal pleiotropy, in which the genetic instruments have effects on the outcome other than through effects on the exposure. MR-Egger, GSMR, weighted median and MR-RAPS are able to provide valid MR estimates under pleiotropy subject to certain assumptions (see ref (30) and supplementary text).

Heterogeneity among the MR estimates across individual SNPs may indicate problems related to violation of instrumental variable assumptions. One of the most notable problems is that one or more SNPs may be showing horizontal pleiotropy (30; 31). The Cochran’s Q statistic and the MR-PRESSO global test (32) were employed to test for heterogeneity for nominally significant MR findings.

#### Interpretation of effect sizes from MR

Regarding the effect sizes of causal associations, if the exposures were binary, the regression coefficients (beta) from MR may be roughly interpreted as average change in the outcome (per SD increase in normalized *ACE2* expression levels) per 2.72-fold increase in the prevalence of the exposure (37). For continuous exposures, the MR estimates are average changes in outcome per unit increase of exposure (see Table S2a for the units).

#### Plasma/serum proteins as exposure and further analysis

In addition to MR analysis on individual plasma/serum proteins, we also performed pathway analysis by ClueGO(33). Hypergeometric tests were conducted on the top-ranked proteins (with p<0.05). As an exploratory analysis, we also searched for drugs with targets overlapping with the top-ranked proteins. Drug targets were defined based on the DrugBank database. Our aim is to uncover drug candidates leading to alteration of *ACE2* expression, which may be therapeutically relevant.

#### Multiple testing correction

We employed a false discovery rate (FDR) approach to multiple testing correction. It controls the expected proportion of false positives among the hypothesis declared significant. FDR is also valid under positive dependency of tests (34).

The FDR in fact depends on the overall fraction of truly null hypothesis, or π_0_. It can also be considered as the prior probability that a null hypothesis is true. In reality, π_0_ may vary for different subgroups of hypotheses. For instance, in our analyses, one may expect different π_0_ for diseases/exposures of different kinds. Previous studies (see Table S1) suggested that some chronic disease patients are more likely affected by the infection. To address the above problem, we adopted an FDR control procedure that accounts for varying prior probabilities of association (i.e. different π_0_) among different types of hypotheses. The procedure is ‘objective’ in the sense that it estimates π_0_ based on the data automatically, without the need to specify π_0_ by the researcher. We employed the methodology ‘FDR regression’ proposed in (35), and the R program by the author[FDRreg(ver 0.2)]. In brief, we divided our hypothesis based on the type of exposure/disease (e.g. respiratory, cardiovascular diseases etc.). These categories served as predictors or covariates, which can be used as input by FDRreg in a regression to estimate the π_0_ of each hypothesis test. We also computed the significance of each predictor; it indicates which categories predicted non-null associations better than chance. For input into FDRreg, we took the results from MR-IVW unless the Egger intercept has p<0.05.

## Results

Please note that all supplementary tables are available at https://drive.google.com/open?id=1XQUn7go8Ycz3V1MdeJbzO81erzkAGPib

### MR analysis for diseases and clinically relevant traits

MR results are presented in Tables 1 and 2 (full results shown in Tables S3 and S4). Traits were shown in main tables if MR-IVW or MR-Egger showed nominally significant (p<0.05) results, and if the number of instrument SNPs>=3 (such that pleiotropy can be assessed and results are more informative).

**Table 1.** Overall MR analysis results achieving nominal significance (p<0.05), with diseases/traits as exposure and *ACE2* lung expression as outcome.

| id | trait | nsnps | b_<br>IVW | pval_<br>IVW | b_<br>Egger | pval_<br>Egger | Egger_<br>intercept | pval_<br>intercept | b_<br>median | pval_<br>median | b_<br>gsmr | pval_<br>gsmr | FDR | pval_het_<br>IVW | pval_het_<br>Egger | P_global_<br>PRESSO |
| --- | --- | --- | --- | --- | --- | --- | --- | --- | --- | --- | --- | --- | --- | --- | --- | --- |
| *Diseases as exposure* |  |  |  |  |  |  |  |  |  |  |  |  |  |  |  |  |
| **<u>Diabetes-related</u>** |  |  |  |  |  |  |  |  |  |  |  |  |  |  |  |  |
| ukb-b-10753 | Diabetes diagnosed by doctor | 58 | 0.175 | 0.015 | 0.002 | 0.988 | 0.020 | 0.225 | 0.146 | 0.183 | 0.171 | 0.017 | **0.055** | 0.350 | 0.379 | 0.304 |
| ukb-b-12948 | Non-cancer illness code,<br>self-reported: diabetes | 49 | 0.162 | 0.034 | 0.203 | 0.255 | -0.005 | 0.797 | 0.126 | 0.280 | 0.158 | 0.034 | **0.067** | 0.322 | 0.288 | 0.306 |
| ukb-b-10694 | Diagnoses - secondary ICD10:<br>E10.9 Type I Diabetes mellitus<br>without complications | 3 | 0.101 | 0.042 | 0.184 | 0.428 | -0.052 | 0.654 | 0.110 | 0.033 | - | - | **0.071** | 0.851 | 0.659 | - |
| ieu-a-23 | Type 2 diabetes | 25 | 0.210 | 0.042 | 0.160 | 0.696 | 0.005 | 0.899 | 0.261 | 0.063 | 0.187 | 0.076 | **0.075** | 0.849 | 0.811 | 0.859 |
| ukb-b-8388 | Started insulin within one year<br>diagnosis of diabetes | 7 | 0.076 | 0.031 | 0.077 | 0.352 | -0.001 | 0.991 | 0.084 | 0.041 | - | - | **0.061** | 0.988 | 0.967 | 0.990 |
| **<u>Neoplasms</u>** |  |  |  |  |  |  |  |  |  |  |  |  |  |  |  |  |
| ukb-d-D12 | Diagnoses - main ICD10: D12<br>Benign neoplasm of colon,<br>rectum, anus and anal canal | 12 | -0.286 | 0.015 | -0.058 | 0.904 | -0.031 | 0.621 | -0.314 | 0.047 | -0.331 | 0.007 | 0.930 | 0.865 | 0.815 | 0.873 |
| ukb-d-C3 | Malignant neoplasm of<br>respiratory system and<br>intrathoracic organs | 3 | 0.514 | 0.009 | 0.711 | 0.511 | -0.056 | 0.821 | 0.416 | 0.047 | - | - | 0.509 | 0.066 | 0.022 | - |
| ieu-a-1134 | ER+ Breast cancer (GWAS) | 7 | 0.176 | 0.020 | 0.539 | 0.128 | -0.094 | 0.259 | 0.122 | 0.224 | - | - | 0.272 | 0.547 | 0.645 | 0.497 |
| ieu-a-1013 | Glioma | 3 | 0.200 | 0.037 | 0.438 | 0.659 | -0.071 | 0.800 | 0.205 | 0.081 | - | - | 0.502 | 0.942 | 0.910 | - |
| **<u>Autoimmune disorders</u>** |  |  |  |  |  |  |  |  |  |  |  |  |  |  |  |  |
| ukb-b-18194 | Non-cancer illness code,<br>self-reported: ankylosing<br>spondylitis | 3 | 0.087 | 0.016 | 0.017 | 0.991 | 0.059 | 0.964 | 0.095 | 0.023 | - | - | 0.112 | 0.944 | 0.735 | - |
| ieu-a-32 | Ulcerative colitis | 29 | 0.026 | 0.667 | 0.586 | 0.003 | -0.105 | 0.003 | 0.099 | 0.240 | 0.026 | 0.626 | **0.054** | 0.171 | 0.629 | 0.165 |
| ieu-a-292 | Inflammatory bowel disease | 107 | 0.003 | 0.938 | 0.246 | 0.019 | -0.031 | 0.011 | 0.023 | 0.728 | 0.010 | 0.827 | 0.126 | 0.724 | 0.846 | 0.700 |
| ieu-a-30 | Crohn's disease | 41 | 0.011 | 0.786 | 0.196 | 0.035 | -0.044 | 0.027 | 0.055 | 0.352 | 0.011 | 0.781 | 0.159 | 0.651 | 0.826 | 0.623 |
| ieu-a-31 | Inflammatory bowel disease | 49 | -0.014 | 0.783 | 0.287 | 0.036 | -0.051 | 0.019 | 0.024 | 0.742 | -0.011 | 0.814 | 0.170 | 0.724 | 0.846 | 0.700 |
| **<u>Other diseases</u>** |  |  |  |  |  |  |  |  |  |  |  |  |  |  |  |  |
| ukb-b-17219 | Diagnoses - secondary ICD10:<br>J45.9 Asthma, unspecified | 19 | 0.264 | 0.035 | 0.877 | 0.056 | -0.064 | 0.152 | 0.295 | 0.092 | 0.269 | 0.035 | 0.500 | 0.769 | 0.826 | 0.770 |
| ukb-b-5115 | Diagnoses - secondary ICD10:<br>Z72.0 Tobacco use | 3 | 0.918 | 0.016 | 0.624 | 0.856 | 0.024 | 0.930 | 0.915 | 0.059 | - | - | 0.510 | 0.415 | 0.187 | - |
| ukb-d-I9 | Diseases of veins, lymphatic<br>vessels and lymph nodes, not<br>elsewhere classified | 14 | -0.269 | 0.024 | -0.138 | 0.656 | -0.017 | 0.645 | -0.174 | 0.265 | -0.264 | 0.028 | 0.934 | 0.886 | 0.858 | 0.900 |
| *Other risk factors or clinically relevant traits as exposure* |  |  |  |  |  |  |  |  |  |  |  |  |  |  |  |  |
| ukb-d-30620_raw | Alanine aminotransferase (U/L) | 91 | 0.047 | 0.007 | 0.081 | 0.041 | -0.012 | 0.341 | 0.069 | 0.015 | 0.051 | 0.004 | 0.223 | 0.717 | 0.716 | 0.722 |
| ukb-d-30070_irnt | Red blood cell (erythrocyte)<br>distribution width (SD) | 225 | 0.243 | 0.020 | 0.349 | 0.066 | -0.004 | 0.500 | 0.208 | 0.191 | 0.239 | 0.023 | 0.555 | 0.524 | 0.514 | 0.537 |
| ukb-d-30220_irnt | Basophill percentage (SD) | 77 | -0.504 | 0.027 | -0.584 | 0.169 | 0.003 | 0.821 | -0.473 | 0.179 | -0.468 | 0.045 | 0.935 | 0.535 | 0.504 | 0.558 |
| ukb-d-30780_raw | LDL direct (mmol/L) | 126 | -0.272 | 0.105 | -0.758 | 0.005 | 0.020 | 0.021 | -0.469 | 0.076 | -0.179 | 0.299 | 0.934 | 0.520 | 0.633 | 0.541 |
| ukb-d-30680_raw | Calcium (mmol/L) | 152 | 0.202 | 0.915 | -10.380 | 0.011 | 0.031 | 0.003 | -1.675 | 0.545 | 0.626 | 0.728 | 0.935 | 0.135 | 0.259 | 0.136 |
| ukb-d-30830_raw | SHBG (mmol/L) | 163 | 0.004 | 0.443 | 0.022 | 0.042 | -0.016 | 0.055 | 0.012 | 0.166 | 0.004 | 0.441 | 0.934 | 0.219 | 0.267 | 0.209 |
| ieu-a-793 | Urate (mg/dl) | 4 | 0.026 | 0.027 | 0.026 | 0.167 | -0.008 | 0.884 | 0.028 | 0.037 | - | - | 0.608 | 0.792 | 0.603 | 0.382 |
| ieu-a-1034 | Height (SD) | 4 | 0.636 | 0.047 | 2.060 | 0.806 | -0.118 | 0.865 | 0.599 | 0.124 | - | - | 0.553 | 0.647 | 0.445 | 0.643 |
| ieu-a-299 | HDL cholesterol (SD) | 84 | 0.084 | 0.515 | 0.563 | 0.022 | -0.026 | 0.020 | 0.065 | 0.758 | 0.086 | 0.508 | 0.581 | 0.575 | 0.714 | 0.592 |

Some items are missing as the number of SNPs is insufficient. Nsnps, number of SNPS. b, beta (causal estimate); pval, p-value. IVW, inverse-variance weighted approach; median, weighted median approach; FDR, false discovery rate, derived from FDR regression; pval\_het, heterogeneity p-value; P\_global\_PRESSO, p-value from the global test of MR-PRESSO (used to assess heterogeneity of MR estimates).
FDR refers to the p-value from MR-IVW (if Egger intercept $p > 0.05$ ) or MR-Egger. Values of FDR below 0.1 are in bold.

**Table 2.** Further MR analysis results for type 2 diabetes based on 2018 Mahajan et al.

| Method | beta | se | pval | Egger<br>Intercept | Intercept<br>pval | n_pleio | nsnps |
| --- | --- | --- | --- | --- | --- | --- | --- |
| MR-IVW | 0.177 | 0.060 | 2.91E-03 | - | - | - | 196 |
| MR-Egger | -0.039 | 0.126 | 0.758 | 0.0159 | 0.0545 | - | 196 |
| GSMR<sup>1</sup> |  |  |  |  |  |  |  |
| $r^2=0.001$ | 0.170 | 0.060 | 4.46E-03 | - | - | 0 | 194 |
| $r^2=0.05$ | 0.140 | 0.035 | 7.12E-05 | | | 0 | 289 |
| $r^2=0.1$ | 0.177 | 0.054 | 9.62E-04 | - | - | 0 | 332 |
| $r^2=0.15$ | 0.146 | 0.027 | 5.93E-08 | | | 0 | 392 |
| $r^2=0.2$ | 0.197 | 0.023 | 9.74E-18 | | | 0 | 448 |
| MR-RAPS<sup>2</sup> | 0.064 | 0.030 | 3.43E-02 | - | - | - | 3737 |

We did not find any evidence of heterogeneity based on Cochran's Q (Heterogeneity $P_{IVW}=0.431/P_{Egger}=0.486$ ) or MR-PRESSO global test ( $p=0.418$ ). We also computed the improvement in model heterogeneity by using MR-Egger over IVW following Rucker's framework. The difference was small and non-significant ( $Q_{IVW}=197.77$ ; $Q_{Egger}=196.06$ ; difference=1.71; $p=0.191$ ).
The exposure GWAS dataset on type 2 diabetes was based on Mahajan et al. (2018). Nature genetics, 50(11), 1505-1513. Instrument SNPs were only selected if they passed genome-wide significance ( $p<5e-8$ ) (except for MR-RAPS). If not otherwise specified, SNPs were clumped at $r^2 = 0.001$ .
1 GSMR can account for correlation among SNPs. We performed GSMR based on SNPs clumped at different $r^2$ clumping thresholds. We consider the association to be more robust if significant results are observed across multiple $r^2$ thresholds
2 MR-RAPS is an MR methodology designed for the inclusion of multiple weak instruments. A more relaxed p-value threshold (0.01) was used for SNP instrument selection.

Overall speaking, 25 traits showed associations with *ACE2* expression at FDR<0.2 and 10 had FDR<0.1 (Table S4). No MR results showed FDR<0.05. There were 68 nominally significant (p<0.05) associations based on MR-IVW and 9 based on MR-Egger. Many significant findings were concentrated on traits related to diabetes.

#### Diabetes-related traits

Remarkably, a number of top-ranked results were related to diabetes. We observed totally five diabetes-related traits that showed nominally significant MR results with FDR<0.1; they were all positively associated with *ACE2* expression. Three are related to diagnosis of diabetes (including both type 1 and 2) in the UKBB. Both doctor-diagnosed diabetes and self-reported diabetes in the UKBB, which presumably composed of mainly type 2 diabetes mellitus (T2DM), were significantly associated with higher *ACE2* expression (MR-IVW p=0.0152 and 0.0343; FDR=0.0547 and 0.0667 respectively). Another finding (id: ieu-a-23) was based on a trans-ethnic meta-analysis on T2DM in 2014 (38) (MR-IVW p=0.0421; FDR=0.0748), which had no overlap with the UKBB sample. The finding of a (nominally) significant result in this dataset can therefore be considered as an independent replication of the UKBB result.

We also observed that starting insulin within one year of diagnosis, which was only assessed within diabetic patients, was causally associated with increased *ACE2* expression (MR-IVW *p*=0.031; FDR=0.061). Early use of insulin may indicate type 1 diabetes (T1DM) as the underlying diagnosis or more severe/late-stage disease for T2DM patients (39). We also observed that as a whole, diabetes-related traits were significantly associated with higher probability of having non-null associations with *ACE2* expression (*p*=0.026; Table S7), based on FDRreg. No evidence of significant directional pleiotropy was observed in the above results (Egger intercept *p*>0.05). We therefore primarily reported the results from MR-IVW, as generally the SE of causal estimates is larger with MR-Egger (36) (hence power is much weaker).

In view of the consistent causal associations with diabetes or related traits, we further searched for GWAS summary statistics that have not been included in the IEU GW AS database. We found another publicly available dataset from the DIAGRAM Consortium, based on a recent meta-analysis of T2DM by Mahajan et al. (28) based on European samples (*N*=898,130). For a more in-depth analysis, we also employed GSMR at various r^2^ thresholds and MR-RAPS in addition to IVW and Egger. The full results are presented in Table 2 (also see Supp. Figures). Reassuringly, with the exception of MR-Egger (which is less powerful (36)), all other methods showed (at least nominally) significant results. For GSMR which accounts for correlated SNPs, it showed significant results consistently across different r^2^ thresholds (lowest *p*=9.74E-18; r^2^ threshold=0.2). While this study (28) has partial overlap with the trans-ethnic analysis in 2014 (38), the consistent associations provide further support to a causal link between diabetes and expression of *ACE2*.

We note that the Egger intercept *p*-value was borderline (p=0.0545), which may raise some concern for pleiotropy. However, we have conducted multiple tests for directional pleiotropy, so false positive findings are possible. The corresponding FDR was 0.999 for this test, if multiple testing was taken into account (573 items). We did not find any evidence of heterogeneity based on Cochran’s Q (Heterogeneity *P*_IVW_=0.431/P_Egger_=0.486) or MR-PRESSO global test (*p*=0.418). To further compare MR-IVW and Egger models, we followed the ‘Rucker framework’ proposed in (30; 40), and computed the improvement in model heterogeneity by using MR-Egger. The difference was small and non-significant (*Q_IVW_*=197.77; *Q_Egger_*=196.06; difference=1.71; *p*=0.191), indicating MR-IVW is a reasonably good fit for the data.

For T2DM or self-reported diabetes from UKBB (which presumably comprised mainly T2DM), the causal estimates ranged from ~0.162 to 0.210. The causal estimate from type 1 diabetes was slightly lower and estimated to be ~0.1006.

#### Other diseases/traits

As shown in Table 1, a number of other diseases/traits also showed (nominally) significant results. Several neoplasms, such as breast and lung cancer, may be associated with increased *ACE2* expression. We also observed that several autoimmune disorders, especially inflammatory bowel diseases may be causally associated with *ACE2* expression. Interestingly, asthma and tobacco use also showed nominal significant associations with higher *ACE2* expression. As for other traits, high alanine aminotransferase (ALT), commonly associated with liver diseases, may be related to elevated *ACE2* expression. Other commonly measured blood measures that may lead to altered *ACE2* expression also included red cell distribution width (often associated with iron-deficiency, folate or B12 deficiency anemia), basophil percentage (inverse relationship), calcium level, urate level, HDL- and LDL-cholesterol (inverse relationship). Note that the FDR is dependent upon the category to which a trait belongs; for example, diabetes-related and autoimmune diseases showed lower FDR, likely because these types of diseases had more significant associations in general. As a tradeoff, other traits/diseases, although having nominally significant results, may have higher FDR. FDR provides an additional reference to guide prioritization of the findings; however, FDR estimation is subject to variability and should not be considered as an absolute guide. Other traits with at least nominal significance may still be worthy of further studies especially if supported by clinical observation or other evidence.

For traits showing nominally significant findings (Table 1), we have also performed other additional analyses. We do *not* observe significant heterogeneity in MR estimates across SNPs (by IVW/Egger) for most traits, except one related to lung cancer (ukb-d-C3). The MR-PRESSO global test was also non-significant for all traits, supporting a lack of heterogeneity. This lack of heterogeneity suggests that substantial horizontal pleiotropy is not very likely. The weighted median estimator supports associations for a subset of traits, including three diabetes-related traits (ukb-b-10694, ieu-a-23, ukb-b-8388). The GSMR method, which removes pleiotropic outliers, is generally consistent with IVW findings (SNPs clumped at r^2^=0.001 for both GSMR/IVW).

### MR results with plasma/serum proteins as exposure

Full results are shown in Table S3 and S4 and the enriched pathways are shown in Table 3 and Table S5. Since a large number of proteins are involved, we only highlight a few top pathways here. Some of top pathways include cytokine and cytokine receptor interaction, VEGFA-VEGF2 signaling pathway, JAS-STAT signaling pathway etc. Table 4 and S6 show the list of drugs whose targets overlap with the top-ranked proteins. Note that the tables do *not* explicitly discern the direction of effects of the drugs. A few drugs target more than one protein. If they are ranked by the number of proteins targeted, the top drugs are fostamatinib, copper, zinc and zonisamide, which target >=3 proteins.

**Table 3.** Top 10 enriched pathways for (nominally) significant proteins in MR analysis.

| GOID | GOTerm | Ontology Source | Term PVal | Term Pval (Bonf corrected) | Associated Genes |
| --- | --- | --- | --- | --- | --- |
| KEGG:04060 | Cytokine-cytokine receptor interaction | KEGG_27.02.2019 | 1.82E-06 | 7.29E-05 | [*CCL25*, *CTF1*, *CX3CL1*, *CXCL12*, *IL15RA*, *IL22*, *IL34*, *IL37*, *LTA*, *LTBR*, *OSM*, *TNFSF4*, *TNFSF8*] |
| WP:3888 | VEGFA-VEGFR2 Signaling Pathway | WikiPathways_27.02.2019 | 7.79E-06 | 3.11E-04 | [*ACACB*, *BCL2L1*, *CFL1*, *EEA1*, *F3*, *IGFBP7*, *JAG1*, *KDR*, *PIK3CA*, *PTPN1*, *TXNIP*] |
| WP:254 | Apoptosis | WikiPathways_27.02.2019 | 1.07E-04 | 4.29E-03 | [*BCL2L1*, *BIRC5*, *DIABLO*, *IGF1*, *LTA*, *MCL1*] |
| WP:3614 | Photodynamic therapy-induced HIF-1 survival signaling | WikiPathways_27.02.2019 | 2.99E-04 | 1.19E-02 | [*BCL2L1*, *BIRC5*, *HK1*, *MCL1*] |
| R-HSA:399954 | Sema3A PAK dependent Axon repulsion | REACTOME_Pathways_27.02.2019 | 3.40E-04 | 1.36E-02 | [*CFL1*, *PAK3*, *PLXNA1*] |
| R-HSA:2173782 | Binding and Uptake of Ligands by Scavenger Receptors | REACTOME_Pathways_27.02.2019 | 4.89E-04 | 1.96E-02 | [*FTH1*, *HP*, *STAB1*, *STAB2*] |
| KEGG:04630 | JAK-STAT signaling pathway | KEGG_27.02.2019 | 5.61E-04 | 2.24E-02 | [*BCL2L1*, *CTF1*, *IL15RA*, *IL22*, *MCL1*, *OSM*, *PIK3CA*] |
| KEGG:04672 | Intestinal immune network for IgA production | KEGG_27.02.2019 | 8.84E-04 | 3.53E-02 | [*CCL25*, *CXCL12*, *IL15RA*, *LTBR*] |
| WP:3657 | Hematopoietic Stem Cell Gene Regulation by GABP alpha/beta Complex | WikiPathways_27.02.2019 | 9.00E-04 | 3.60E-02 | [*BCL2L1*, *FLT3*, *MCL1*] |
| WP:3872 | Regulation of Apoptosis by Parathyroid Hormone-related Protein | WikiPathways_27.02.2019 | 0.00090 | 0.03602 | [*BCL2L1*, *MCL1*, *PIK3CG*] |

Pval, p-value; Bonf, Bonferroni correction.

**Table 4.**
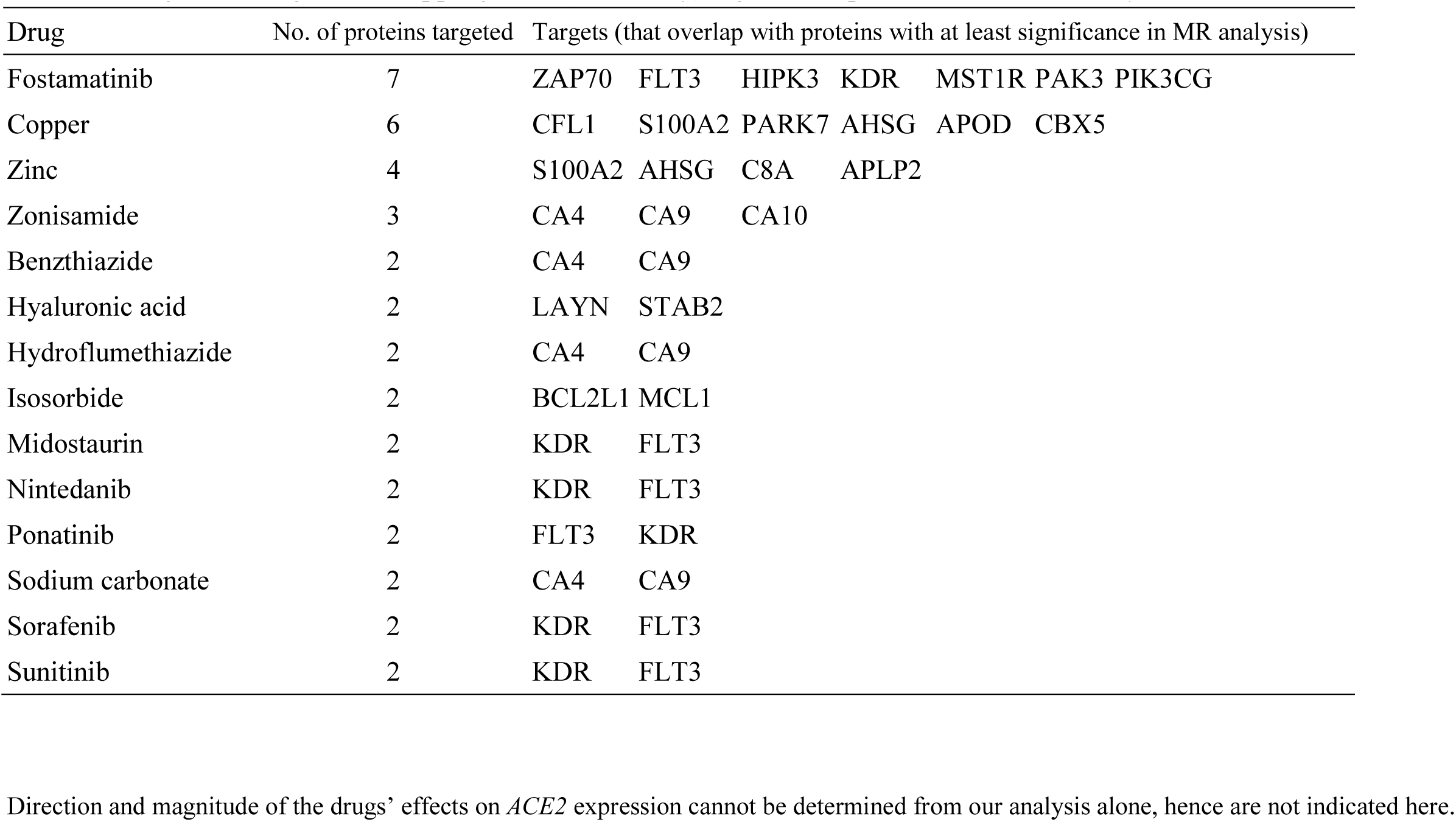
Drugs with targets overlapping with (nominally) significant proteins from MR analysis

## Discussions

In this study we have employed Mendelian Randomization (MR) to uncover diseases/traits that may be causally linked to *ACE2* expression in the lung, which in turn may influence susceptibility to the infection. MR is a relatively well-established technique in evaluating causal relationships, and the wide availability of GWAS data enables many different exposures to be studied in the same time.

### Diseases/traits causally linked to ACE2 expression

From our analysis, the most consistent finding was the tentative causal link between diabetes (and related traits) with *ACE2* expression, which was supported by multiple datasets and different analytic approaches. Other results were more tentative, but may be worthy of further studies. For example, several neoplasms (e.g. breast and lung cancers) and autoimmune diseases, elevated ALT, asthma and smoking all showed nominally significant and positive associations with *ACE2* expression.

Some of these findings were supported by previous studies. A number of COVID-19 cases (~5.4% from Table S1) were comorbid with diabetes mellitus (DM). This proportion is only a rough estimate since mild or asymptomatic cases may remain undetected. Notably, DM has been reported to be associated with poorer outcomes among infected patients (41). Similarly, DM was also common in patients infected with MERS-CoV (42; 43). Kulcsar *et al*. built a mouse model susceptible to MERS-CoV infection and induced T2DM using a high-fat diet. They found that, if affected by the virus, these diabetic mice suffered from a prolonged phase of disease and delayed recovery, possibly due to a dysregulated immune response (44). Regarding comorbidity with cancers, Liang *et al*. recently carried out a nationwide analysis of 1,590 patients with confirmed COVID-19 and suggested that cancer patients have higher infection and complication risks than those without (45).

We highlight a few research directions of interest, if our findings are confirmed in future studies. For example, as far as treatment is concerned, if certain conditions (e.g. diabetes) increase susceptibility to infection or severe infections via ACE2, drugs targeting this gene/protein may be particularly useful for this patient subgroup. For example, human recombinant ACE2 has been proposed as a treatment and is under clinical trial (46; 47). It will be interesting to see if the drug may be more beneficial for DM patients. More generally speaking, if DM is causally linked to elevated *ACE2* and potentially increased susceptibility to infection, then anti-diabetic drugs or improved glycemic control may ameliorate the process. Interestingly, a recent work highlighted metformin as one of the top repositioning candidates for COVID-19, based on a different mechanism as an MRC1 inhibitor (48). From a public health perspective, identification of at-risk populations may guide prevention strategies, e.g. prioritization of groups to receive vaccination. Nevertheless, all the above require substantial additional research before clinical applications.

### On ACE2 expression and pulmonary complications

As discussed above, increased expression of *ACE2* appears to correlate with susceptibility to SARS-CoV and SARS-CoV-2 infection. Nevertheless, the consequences of altered *ACE2* expression on pulmonary complications may be rather complex. Kuba *et al*. reported that SARS-CoV down-modulated *ACE2* expression (7), which may lead to heightened risks of acute lung injury (ALI). Another study (49) suggested ACE2 may protect against ALI by blocking the renin-angiotensin pathway. However, whether the same applies to SARS-CoV-2 is unknown. If this is the case, one may hypothesize that for *unaffected* individuals or those without (or with minimal) lung involvement yet, *lower ACE2* pulmonary expression may be beneficial in reducing susceptibility to more sustained infection by reducing viral entry. However, for patients with severe lung involvement or at risk of ALI, *higher ACE2* expression may prevent acute respiratory failure. Therefore, it may be clinically relevant to identify both risk factors/drugs leading to increased and decreased *ACE2* expression. Further studies are warranted to clarify the role of ACE2 in COVID-19 and related complications.

Another related controversy concerns the use of ACE inhibitors (ACEI) and angiotensin II receptor blockers (ARB) (50; 51), although the present study does not directly address this issue. There is some evidence that ACEI/ARB may upregulate *ACE2* expression in the heart (52), kidney (53) and aorta (54) in animal models, however how these drugs affect pulmonary *ACE2* levels in humans is still unclear (55). In addition, it is possible that patients’ other underlying conditions may affect *ACE2* expression. It is worthy to further investigate how ACEI/ARB together with other chronic conditions affects the risks and severity of infection.

### Highlight of tentative repositioning candidates based on blood proteins potentially linked to ACE2 expression

The drugs we highlighted in this study may help researchers to *prioritize* repositioning candidates for further studies, given the huge cost and long time in developing a brand-new drug. Nevertheless, the overall direction and magnitude of effect of each drug could not be determined from our analysis alone, hence further studies are required. Here we briefly highlight a few top candidates. *Fostamatinib* targets the largest number (seven) of proteins potentially linked to *ACE2* expression. According to DrugBank, it serves as an inhibitor for all these proteins, and all were linked to elevated *ACE2* expression except one. Interestingly, a recent computational repositioning study (56; 57) identified baricitinib, a JAK 1/2 and AAK1 inhibitor approved for rheumatoid arthritis (RA) as a top candidate. Fostamatinib is a spleen tyrosine kinase inhibitor but also inhibits JAK 1/2 and AAK1 (from DrugBank) (58) and can be used to treat RA (59). JAK-STAT signaling was also among the top 10 pathways enriched for top proteins affecting *ACE2* expression. Interestingly, fostamatinib was reported to be effective for type 1 diabetes (60). Another candidate highlighted in (56), sunitinib, was also top-listed by our analysis. *Zinc* is also a top-listed candidate, which was reported to reduce risks of lower respiratory tract infections (61), but the evidence is not firm. Interestingly, a study in rat tissues showed reduction of ACE2 activity by zinc (62). Zinc and zinc-ionophores may inhibit SARS-CoV as shown in experimental studies (63). Zinc was recently suggested for clinical trials for COVID-19, although there is no clinical evidence yet (clinicalTrials.gov NCT04342728, NCT04326725, NCT04351490, ref (64)). As for the enriched pathways for top-ranked proteins affecting *ACE2* expression, they are discussed in Supplementary Text.

### Limitations

We wish to emphasize that we consider this work as largely an *exploratory* rather than confirmatory study. As such, the findings might not be immediately applicable clinically. Our main purpose is to *prioritize* diseases, traits or proteins with potential causal links with *ACE2* expression. There are several limitations in our analysis. A major limitation is that the sample size for GTEx is relatively modest (*N* = 515), which limits the power of MR analysis. However, to our knowledge, GTEx is one of the largest databases with both genotype and expression data. We note that many associations were relatively modest, with no results showing FDR<0.05, although 25 had FDR<0.2. On the other hand, we examined the consistency of the observed associations across different datasets, and considered those supported by more than one set of data (e.g. diabetes-related traits) as relatively more robust, similar to the approach in (65). However, our findings will require further support by further studies. Besides, some results could be false negatives owing to limited power. Besides, while most GWAS were based on predominantly European samples, subjects of other ethnicities were included in some samples. It is possible for genetic associations to differ across ethnicities, which may affect the causal estimates of MR, for example if some SNP-exposure or SNP-outcome associations are stronger in one ethnic group than another. Apart from the above, this study does *not* address what factors may aggravate or ameliorate *CoV-induced* changes in ACE2 levels. Also, we studied *ACE2* mRNA expression as the outcome; associations of the reported traits with protein expression levels remain to be investigated.

Finally, from a methodological point of view, we have employed MR in a different manner from most other studies. Usually MR is used to identify causal risk factors with a disease as the outcome, for which GWAS data is available. Here we presented a novel analytic approach: we made use of existing knowledge of a key receptor of an infectious agent to uncover causal risk factors and repositioning candidates. This analytic framework may also be applied to other diseases, especially when a target can be identified but genomic data for the disease is limited, or if one is interested in the underlying disease mechanism of the risk factor.

## Conclusions

Notwithstanding the limitations, we have identified several diseases and traits which may be causally related to *ACE2* expression the lung, which in turn may mediate susceptibility to SARS-CoV-2 infection. In addition, our proteome-wide MR analysis revealed proteins that could lead to changes in *ACE2* expression. Subsequent drug repositioning analysis highlighted several candidates that may warrant further investigations. We stress that most of the findings require validation in further studies, especially the part on repositioning. Nevertheless, we believe this work is of value in view of the urgency to address the outbreak of COVID-19.

## Data Availability

Data are available from the IEU-GWAS and GTEx databases. GWAS summary statistics for diabetes is also available from the DIAGRAM Consortium website.

## Acknowledgements

We would like to thank Prof. Stephen Tsui for computing support. This study was partially supported by the Lo Kwee Seong Biomedical Research Fund, an NSFC grant (81971706) and a Chinese University of Hong Kong Direct Grant. We also thank Mr. Carlos Chau for assistance in part of the analysis.

## Author contributions

Conceived and designed the study: HCS (lead), with input from SR. Supervised the study: HCS. Data analysis: HCS (lead), SR, AL. Data interpretation: HCS, SR, AL. Drafted the manuscript: HCS (lead), with input from AL and SR.

## Conflicts of interest

The authors declare no conflict of interest.

## Notes

### Competing Interest Statement

The authors have declared no competing interest.

### Funding Statement

This study was partially supported by the Lo Kwee Seong Biomedical Research Fund, an NSFC grant and a Chinese University of Hong Kong Direct Grant.

